# Use of convalescent plasma in patients with coronavirus disease (Covid-19): Systematic review and meta-analysis

**DOI:** 10.1101/2021.02.14.20246454

**Authors:** Fernando Tortosa, Gabriela Carrasco, Martin Ragusa, Pedro Haluska, Ariel Izcovich

**Author notes:** Correspondence. Fernando Tortosa Moreno 601 SanCarlos de Bariloche, Rio Negro, Argentina CP 8400. All authors declare that they have no conflicts of interest. We do not have external sources of financing: this study was carried out thanks to the support of the Ministry of Health of the province of Rio Negro.

## Abstract

**Objetives:** To assess the effects of convalescent plasma treatment in patients with coronavirus disease (COVID-19).

**Study design:** Systematic review and Meta-analysis

**Data sources:** A systematic search was carried out on the L · OVE (Living OVerview of Evidence) platform for COVID-19 until October 31, 2020

**Study selection:** Randomized clinical trials in which people with probable or confirmed COVID-19 were randomized to drug treatment, standard care, or placebo. Pairs of reviewers independently screened potentially eligible articles.

**Methods:** The PRISMA guidelines were followed for conducting a systematic review and meta-analysis. The risk of bias of the included studies was assessed using the Cochrane risk of bias tool 2.0, and the certainty of the evidence using the recommendation assessment, development and evaluation (GRADE) approach. For each outcome, the interventions were classified into groups, from most to least beneficial or harmful.

**Results:** We identified 10 RCTs (randomized controlled trials) involving 11854 patients in which convalescent plasma was compared with standard of care or other treatments. The results of five RCTs that evaluated the use of convalescent plasma in patients with COVID-19 did not show significant differences in the effect on mortality and the need for invasive mechanical ventilation.

**Conclusions:** Current evidence is insufficient to recommend the use of convalescent plasma in the treatment of moderate or severe COVID-19.

**Contribution of the authors:** 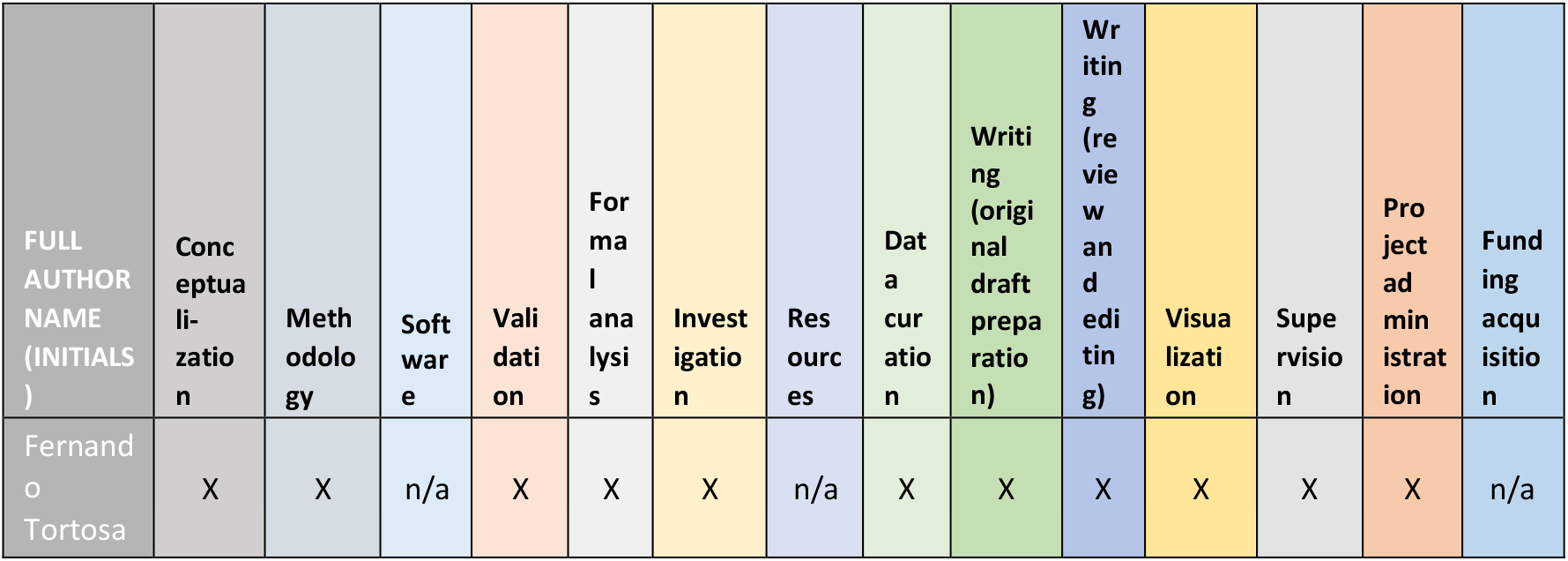

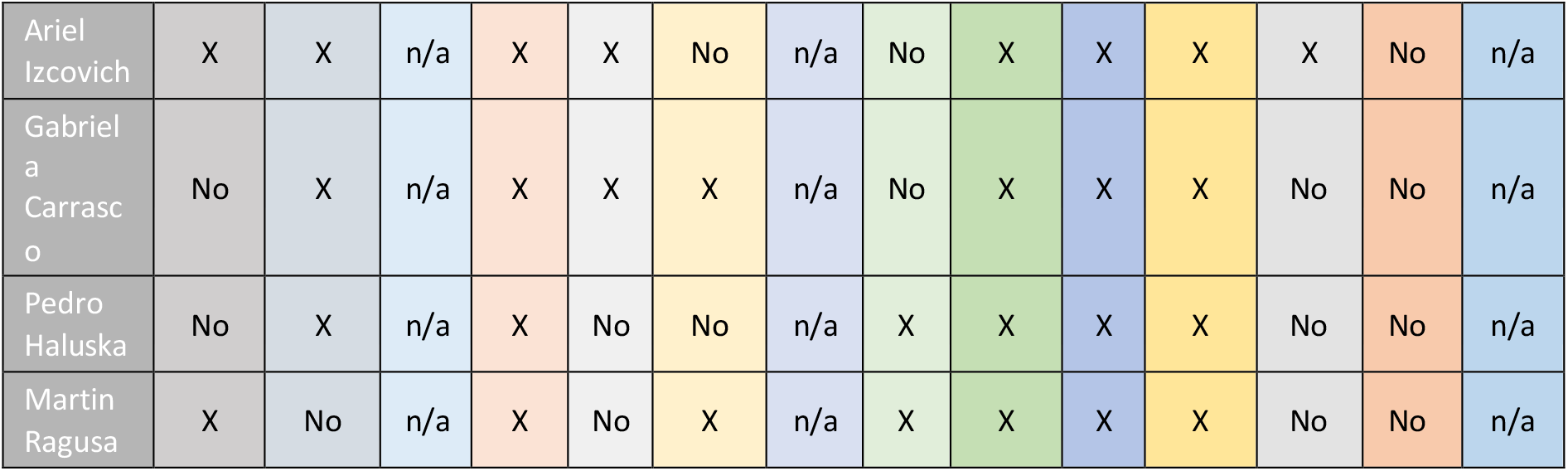

## Introduction

Patients who have recovered from infectious diseases have antibodies in their blood that protect them from future diseases caused by the same infectious agent. These antibodies can be obtained from blood plasma, which in this context is called convalescent plasma.

The transfusion of convalescent plasma to a person with a viral infection could neutralize the pathogenic microorganism that affects them and, thus, give that person time to initiate an active immune response, that is, generated by their own immune system.

Some believe that this therapy played a fundamental role at a time when we did not have effective vaccines or medications for most diseases. The antecedent of successful use in our country arises from the treatment of Argentine Hemorrhagic Fever [1] [2]. As we are in precisely that situation today in the face of COVID-19, interest in using it has resurfaced. But if we had an efficient vaccine or drug today, the option of using convalescent plasma would probably not be considered to use convalescent plasma, because there are practical difficulties in obtaining it and its availability, consequently, is limited [3]. The central thing to consider could be an alternative for the treatment of COVID-19, while scientists discover a better option, however, the certainty that it could work in other situations is also very low.

We conducted a systematic review to summarize the available evidence regarding the use of convalescent plasma in patients with coronavirus disease (COVID-19).

## Methods

This study was conducted following the PRISMA guidelines (prefered reported items in systematic reviews and meta-analysis) for conducting and publishing systematic reviews [4].

The protocol was registered in PROSPERO with registration number CRD42020226735 “Use of convalescent plasma in patients with coronavirus disease (Covid-19): systematic review and meta-analysis”.

### objective

To assess the effects of convalescent plasma treatment in patients with coronavirus disease (COVID-19) compared to standard treatment or placebo.

### Clinical question

In people with coronavirus disease (COVID-19), should convalescent plasma be used compared to standard treatment alone?

### PICO question

Population People with coronavirus disease (COVID-19)

Intervention Convalescent plasma plus standard treatment

Comparison Standard Treatment or Placebo

Outcomes Mortality, admission to AVM (mechanical ventilatory assistance), serious adverse events, length of hospitalization, time of mechanical ventilation, length of stay in the ICU (Intensive Care Unit)

### Design

Systematic review and meta-analysis

### Data sources

Systematic searches were carried out on the L · OVE (Living OVerview of Evidence) platform for COVID-19, a system that maps PICO questions to a repository developed by the Epistemonikos Foundation, this repository is continuously updated by searching electronic databases, server pre-printing, test logs and other resources relevant to COVID-19. The electronic databases PubMed, LiLacs, Cochrane Library and the clinicaltrials.gov register of clinical trials were also searched in duplicate. For more information see Supplementary Information Annex.

No search restrictions were imposed on electronic databases. The last search date was carried out on November 1, 2020.

### Study selection

Randomized controlled trials (RCTs) were selected for this therapeutic pharmacological intervention (without age restriction) with confirmed COVID-19 and trials comparing this intervention head-to-head or with no intervention or placebo, providing evidence on critical or important outcomes, were included. (mortality, mechanical ventilation, symptoms resolution or clinical improvement, and serious adverse events).

Two review authors independently screened all titles and abstracts, followed by the full texts of the trials that were identified as potentially eligible. A third reviewer resolved the conflicts.

### Data collection

For each eligible trial, pairs of reviewers independently extracted data using a data extraction form.

Reviewers collected information on trial characteristics (trial registration, publication status, study status, design), participant characteristics, and outcomes of interest. The reviewers resolved discrepancies by discussion and, where necessary, with the intervention of a third reviewer.

### Risk of bias of included studies

An assessment of risk of bias was applied to RCTs based on randomization, allocation, concealment, blinding or other biases relevant to sources of risk of bias. For each eligible trial and outcome, the reviewers used a review using the Cochrane Risk of Bias in RCTs (RoB 2.0) [5] to rate trials in the following domains: bias arising from the randomisation process; bias due to deviations from the planned intervention; bias due to missing outcome data; bias in the measurement of the outcome; bias in the selection of outcome reports (including deviations from the recorded protocol) and bias arising from early termination for the benefit of the intervention. Trials were rated at high risk of bias overall if one or more domains were rated as ‘Some concerns or likely high risk of bias’ or as ‘high risk of bias’, and as low risk of bias overall, if all domains were rated as “some concerns, probably low risk of bias”. The reviewers resolved discrepancies by discussion and when not possible they were resolved by a third party.

### Effect measures and statistical analysis

We summarize the effect of interventions on dichotomous outcomes using relative risks (RR) and corresponding 95% confidence intervals (CI).

For any meta-analytic pooling, whenever the data allowed, all studies were pooled. The relative effect was calculated through a relative risk meta-analysis using the Mantel and Haenszel method with a random effect model. The absolute effect was estimated from the relative risk and the risk observed in control groups of the included studies.

We performed meta-analysis of direct comparisons and the rest of the calculations using the “meta” package in RStudio Version 1.3.1093.14. [6]

To assess the absolute effects of the interventions, related effects applied to baseline risks (risks without intervention), mortality and baseline risks of mechanical ventilation were extracted from the ISARIC cohort (https://isaric.tghn.org/) and for adverse events and the resolution and / or improvement of symptoms, we used the risk observed in the control or standard treatment groups of the included RCTs. [7]

### Certainty of the evidence

We assessed the certainty of the evidence by grading the recommendations assessment, development approach and assessment (GRADE) [8]. Two methodologists experienced in using GRADE scored each domain for each comparison separately and resolved discrepancies by consensus. We rated the certainty for each comparison and outcome as high, moderate, low, or very low, based on considerations of risk of bias, inconsistency, indirect, publication bias, and imprecision.

Sensitivity, subgroup and publication bias analysis

The risk of bias according to current standards for pharmacological interventions in COVID-19 and the quality of the infused plasma were considered as potential subgroup effects or effect modifiers, based on the concentration of neutralizing antibodies required in the standards set in the clinical trials.

The presence of publication bias was analyzed by performing a funnel plot and using the Eggers test statistic [9].

### Sample power

A sample size calculation was performed to determine whether the total number of patients included in the meta-analysis was less than the required number of patients to achieve sample power and to determine if the optimal information size (TOI) was met for a power 80%. (d = 0.10, RR, k = 6, n1 = 500, n2 = 500, p = 0.05, heterogeneity = “moderate”) [10].

## Results

Five RCTs [11] [12] [13] [14] [15] were identified that included 734 patients in which convalescent plasma was compared with standard treatment or other treatments.

The characteristics of the included studies can be seen in Table 1. Agarwal et al conducted the largest study to date that included 235 patients in the intervention arm and 229 in control.

All studies included severe and moderate patients. Mortality in the control arms ranged from 10% to 25.6%. Convalescent plasma was administered in one or two infusions to symptomatic patients in all cases. The risk of bias of the studies was low overall for death or admission to AVM and high for therest of the outcomes. This was due to problems in randomization and blinding in most studies,the presence of multiple co-interventions and the reporting of results. See Chart 2 for risk of bias.

**Graph 2.**
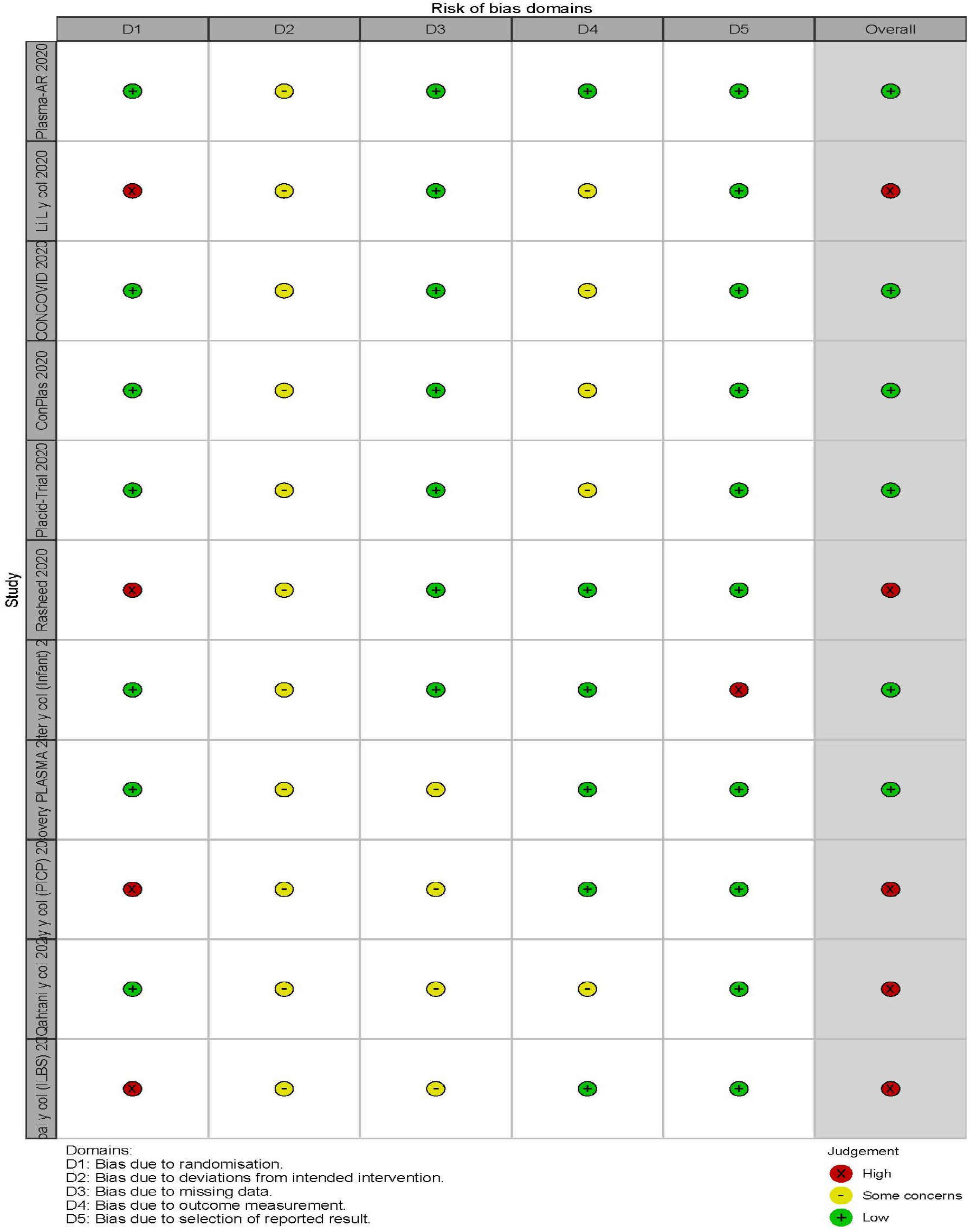
Risk of bias of the included studies

With 10 studies included and more than 5000 patients per branch, 100% of the sample power was reached with optimal information size determination (TOI).

### Outcome mortality

The use of convalescent plasma has no impact on mortality of patients interned with COVID-19, RR 1.02 (95% CI: 0.94 to 1.12); RD 0.2% more (0.6 minus 1 more); HIGH Certainty ⨁ ⨁ ⨁ ⨁.. see Chart 3 and 3a Forest plot

**Chart 3.**
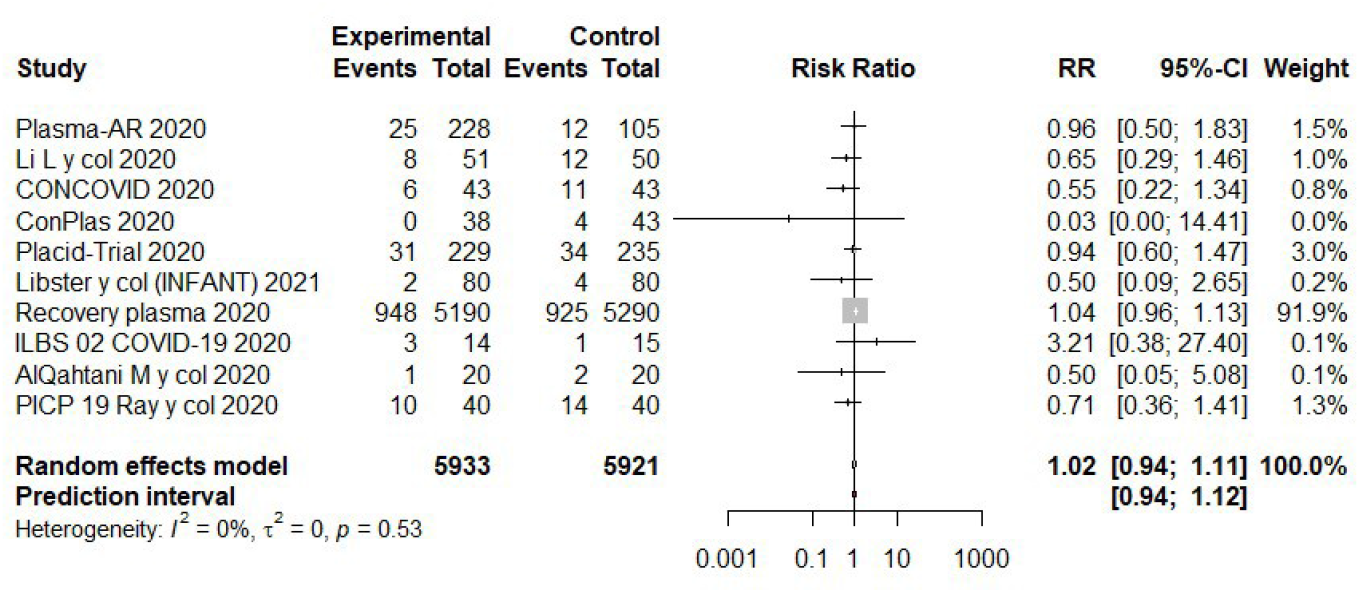
Forest plot. **Effect of convalescent plasma use, Outcome: Mortality at 30 days, Randomized controlled studies. Chart 3a Forest plot subgroup effects: Studies with high risk of bias vs low risk of bias**.

**Chart 3.**
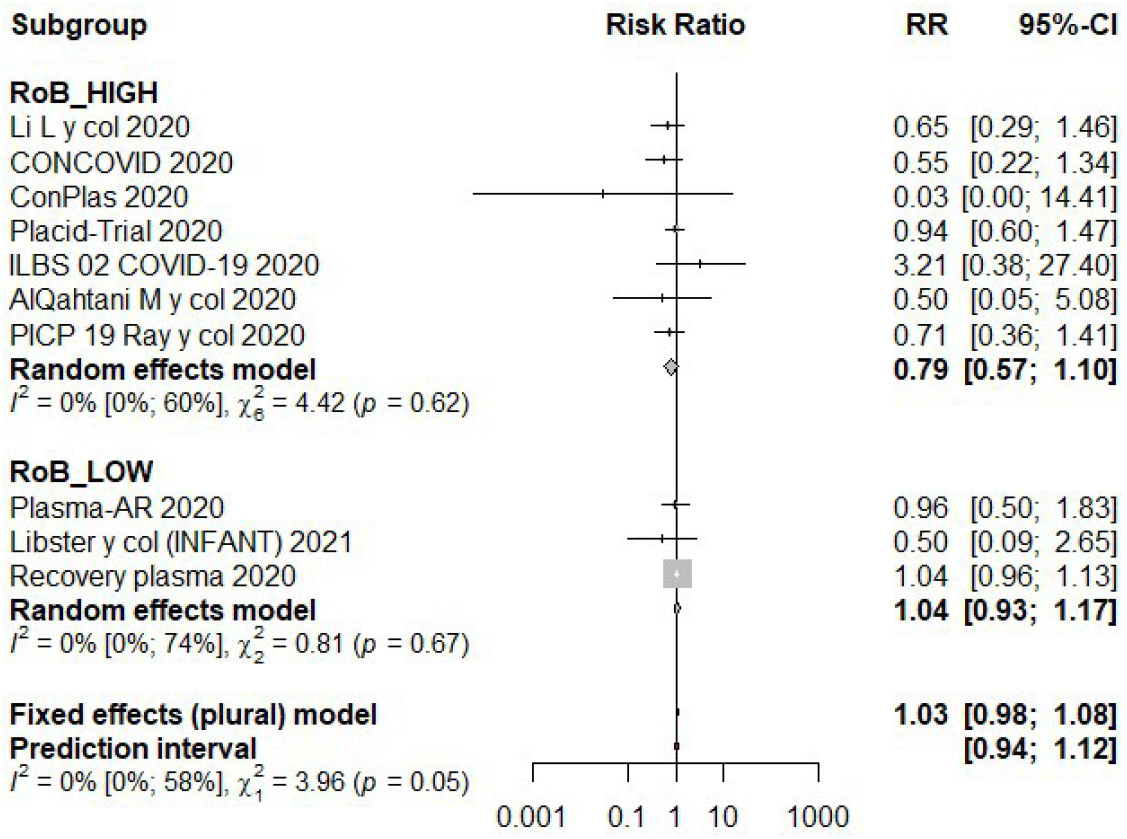
Forest plot. **Effect of convalescent plasma use, Outcome: Mortality at 30 days, Randomized controlled studies. Subgroup analysis: Studies with high risk of bias and/or low antibody tidy vs low risk of bias and high antibody tidys**.

**Convalescent plasma could produce a marginal increase in admission to mechanical ventilation, RR 1**.**17 (95% CI 0**.**80 to 1**.**70); RD 2% (95% CI −2% to 8%);**

**Low certainty ⨁ ⨁ ◯◯. see Graph 4 Forest plot**

**Graph 4.**
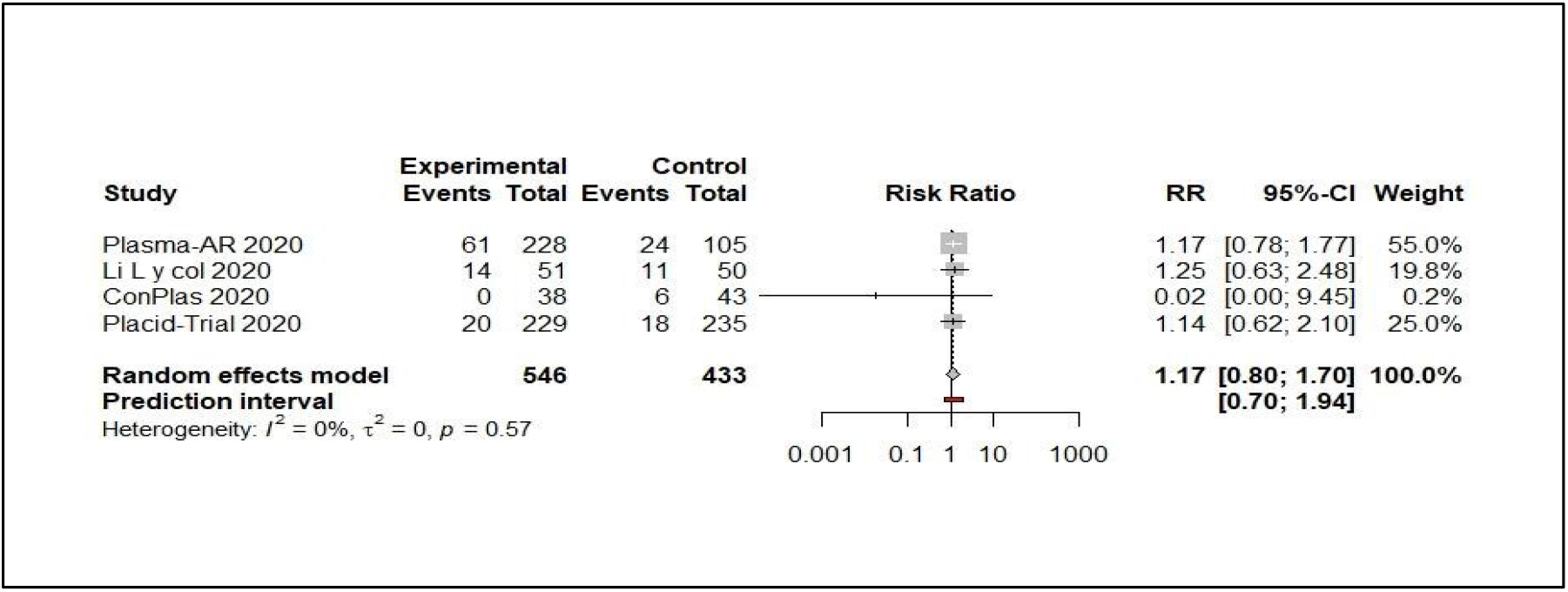
Forest plot. **Effect of the use of convalescent plasma, Outcome: Admission to mechanical ventilatory assistance (MVA), Subgroup Randomized controlled studies**

**Outcome serious adverse events**

**Convalescent plasma could produce a marginal increase in serious adverse events, RR 1**.**27 (95% CI 0**.**72 to 2**.**24); RD 1**.**4% (95% CI: −1**.**6% to 6**.**7%); Low certainty ⨁ ⨁**

**◯◯**.

**Graph 5.**
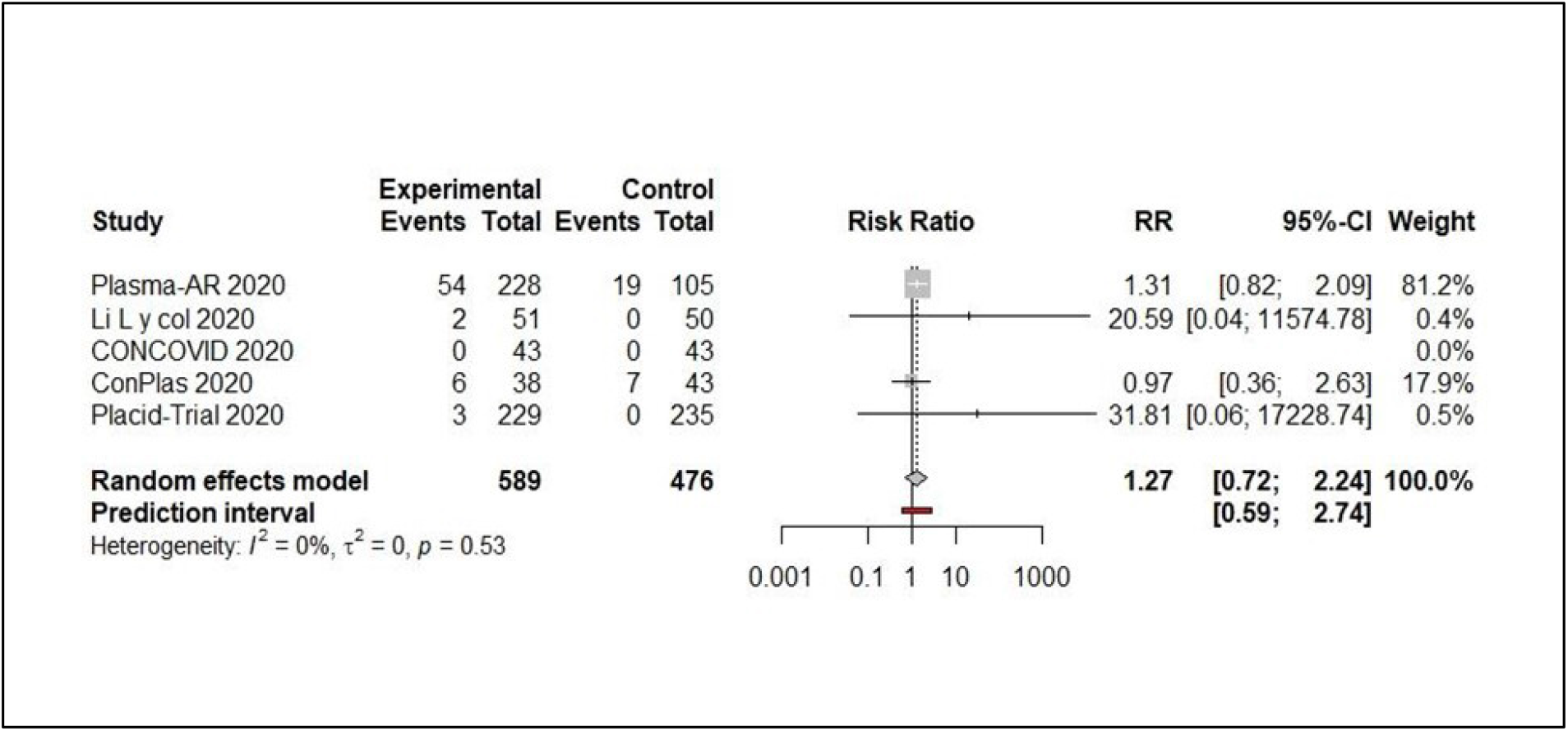
Forest plot. **Convalescent plasma vs. SOC for the treatment of Covid-19.** **Outcome: serious adverse events**

## Discussion

Evidence from cohorts [21] [22] [23] [24] where convalescent plasma was openly infused to large numbers of people provides us with a large amount of data, but this evidence is nonetheless very low. certainty.

One of the observations made on this type of observational study such as the one carried out by the Mayo Clinic [21] is the low occurrence of adverse events (transfusion-related volume overload 0.4%, transfusion-related acute pulmonary injury 0, 22% and severe transfusion allergic reaction 0.06%). However, given the clinical characteristics in the evolution of the coronavirus disease (sometimes clinical and imaging manifestations difficult to distinguish, for example, from a transfusion-related lung injury) and the lack of rigor in the reports of adverse events in this type of registry (or substantially less than that of a clinical trial) is that we prefer to prioritize the certainty of the evidence in adverse events occurring in clinical trials (see evidence profile).

In our study, we found that the use of convalescent plasma does not impact the mortality of patients with moderate to critical illness.

It could be associated in a borderline manner with an increase in admission to mechanical ventilation and in the incidence of serious adverse events. Regarding other less important outcomes such as time to clinical improvement, there is also uncertainty in the effect of the intervention. These statements are based on a low certainty of the evidence, due to the risk of bias in the included studies and unexplained heterogeneity. Subgroup analyzes performed for risk of bias and quality of the infused product (related to antibody concentration) could not explain this heterogeneity. We can hypothesize that the co-interventions, the severity of the patients included through the studies and the timing of the infusion could influence it.

## Conclusion

The results of ten RCTs (randomized controlled trials) that evaluated the use of convalescent plasma in patients with COVID-19 did not show significant differences in the effect on mortality and the need for invasive mechanical ventilation. For some outcomes, the certainty in the evidence is still low and more information is needed from properly designed studies to be able to improve the certainty in the direction and magnitude of the effect.

One of the greatest difficulties with this and other interventions that are used in the treatment of Covid-19, is the form of registration of interventions of unproven efficacy or still without preclinical evidence or in phases I and II of research. This deficit of records, deficient records or not intended to provide reliable data, cause a significant delay in knowledge in this area.

Current evidence shows that the use of convalescent plasma has no effect on critical outcomes in patients with moderate or severe COVID-19. Its early use, in moderate high-risk patients with a product with a very high antibody titer, based on its biological plausibility, has an uncertain benefit and is not feasible to implement, and could negatively impact the equity of the health system.

## Data Availability

We have all data

## Financing sources

We do not have external sources of financing to carry out this study

## References

1. Instituto Nacional de Enfermedades Virales Humanas “Dr. Julio I. Maiztegui” (1997). Fiebre hemorrágica Argentina. Actualización sobre diagnóstico, tratamiento y prevención.

2. Sanchez, J. D. (s. f.). Fiebre Hemorrágica Argentina. Recuperado 23 de junio de 2020, de https://www.paho.org/hq/index.php?option=com_content&view=article&id=8306:2013-fiebre-hemorragica-argentina&Itemid=39845&lang=es

3. Fundación Epistemonikos. (2020, 19 abril). ¿Podría ser efectivo el plasma convaleciente para tratar COVID-19? Recuperado 22 de junio de 2020, de https://es.epistemonikos.cl/2020/04/17/podria-ser-efectivo-el-plasma-convalecientepara-tratar-covid-19/

4. Liberati A, Altman DG, Tetzlaff J, Mulrow C, Gøtzsche PC, Ioannidis JP, Clarke M, Devereaux PJ, Kleijnen J, Moher D. The PRISMA statement for reporting systematic reviews and meta-analyses of studies that evaluate healthcare interventions: explanation and elaboration. BMJ. 2009 Jul 21;339:b2700. doi:10.1136/bmj.b2700. PMID: 19622552; PMCID: PMC2714672.

5. Sterne JAC, Savović J, Page MJ, Elbers RG, Blencowe NS, Boutron I, et al. RoB 2: a revised tool for assessing risk of bias in randomised trials. BMJ. 28 de 2019;366:4898.

6. Balduzzi S, Rücker G, Schwarzer G. How to perform a meta-analysis with R: a practical tutorial. Evid Based Ment Health. noviembre de 2019;22(4):153–60.

7. Hall M, Pritchard M, Dankwa EA, Baillie JK, Carson G, Consortium ISAR and emerging I, et al. ISARIC Clinical Data Report 20 November 2020. medRxiv. 23 de noviembre de 2020;2020.07.17.20155218.

8. Atkins D, Best D, Briss PA, Eccles M, Falck-Ytter Y, Flottorp S, et al. Grading quality of evidence and strength of recommendations. BMJ. 19 de junio de 2004;328(7454):1490.

9. Egger M, Davey Smith G, Schneider M, Minder C. Bias in meta-analysis detected by a simple, graphical test. BMJ. 1997 Sep 13;315(7109):629–34. doi:10.1136/bmj.315.7109.629. PMID: 9310563; PMCID: PMC2127453.

10. Borenstein, M., Hedges, L. V., Higgins, J. P., & Rothstein, H. R. (2011). Introduction to meta-analysis. John Wiley & Sons. (Chapter 29).

11. Avendano-Sola C, Ramos-Martinez A, Munez-Rubio E, Ruiz-Antoran B, Malo de Molina R, Torres F, et al. Convalescent Plasma for COVID-19: A multicenter, randomized clinical trial. medRxiv [Internet]. 2020; Disponible en: https://www.medrxiv.org/content/early/2020/09/29/2020.08.26.20182444

12. Li L, Zhang W, Hu Y, Tong X, Zheng S, Yang J, et al. Effect of Convalescent Plasma Therapy on Time to Clinical Improvement in Patients With Severe and Life-threatening COVID-19. JAMA. de agosto de 2020;324(5):1–11.

13. Gharbharan A, Jordans CCE, GeurtsvanKessel C, den Hollander JG, Karim F, Mollema FPN, et al. Convalescent Plasma for COVID-19. A randomized clinical trial. medRxiv [Internet]. 2020; Disponible en: https://www.medrxiv.org/content/early/2020/07/03/2020.07.01.20139857

14. Simonovich VA, Burgos Pratx LD, Scibona P, Beruto MV, Vallone MG, et al; PlasmAr Study Group. A Randomized Trial of Convalescent Plasma in Covid-19 Severe Pneumonia. N Engl J Med. 2020 Nov 24. doi:10.1056/NEJMoa2031304. Epub ahead of print. PMID: 33232588.

15. Agarwal A, Mukherjee A, Kumar G, Chatterjee P, Bhatnagar T, Malhotra P, et al. Convalescent plasma in the management of moderate covid-19 in adults in India: open label phase II multicentre randomised controlled trial (PLACID Trial). BMJ. 22 de 2020;371:m3939.

16. Ray Y, Paul SR, Bandopadhyay P, D’Rozario R, Sarif J, Lahiri A, et al. Clinical and immunological benefits of convalescent plasma therapy in severe COVID-19: insights from a single center open label randomised control trial. medRxiv. 1 de enero de 2020;2020.11.25.20237883.

17. Libster R, Pérez Marc G, Wappner D, Coviello S, Bianchi A, Braem V, et al. Early High-Titer Plasma Therapy to Prevent Severe Covid-19 in Older Adults. New England Journal of Medicine. 0(0):null.

18. Bajpai M, Kumar S, Maheshwari A, Chhabra K, kale P, Gupta A, et al. Efficacy of Convalescent Plasma Therapy compared to Fresh Frozen Plasma in Severely ill COVID19 Patients: A Pilot Randomized Controlled Trial. medRxiv. 1 de enero de 2020;2020.10.25.20219337.

19. AlQahtani M, Abdulrahman A, Almadani A, Alali SY, Al Zamrooni AM, Hejab AH, et al. Randomized controlled trial of convalescent plasma therapy against standard therapy in patients with severe COVID-19 disease. medRxiv. 1 de enero de 2020;2020.11.02.20224303.

20. RECOVERY trial closes recruitment to convalescent plasma treatment for patients hospitalised with COVID-19 — RECOVERY Trial [Internet]. [citado 30 de enero de 2021]. Disponible en: https://www.recoverytrial.net/news/statement-from-therecovery-trial-chief-investigators-15-january-2021-recovery-trial-closes-recruitmentto-convalescent-plasma-treatment-for-patients-hospitalised-with-covid-19

21. Rasheed AM, Fatak DF, Hashim HA, Maulood MF, Kabah KK, Almusawi YA, Abdulamir AS. The therapeutic potential of convalescent plasma therapy on treating critically-ill COVID-19 patients residing in respiratory care units in hospitals in Baghdad, Iraq. Infez Med. 2020 Sep 1;28(3):357–366. PMID: 32920571.

22. Salazar, E., Christensen, P. A., Graviss, E. A., Nguyen, D. T., Castillo, B., Chen, J., et al. Significantly Decreased Mortality in a Large Cohort of Coronavirus Disease 2019 (COVID-19. Patients Transfused Early with Convalescent Plasma Containing High-Titer AntiSevere Acute Respiratory Syndrome Coronavirus 2 (SARS-CoV-2) Spike Protein IgG. The American journal of pathology, S0002-9440(20)30489-2. Advance online publication. https://doi.org/10.1016/j.ajpath.2020.10.008

23. Joyner MJ, Bruno KA, Klassen SA, Kunze KL, Johnson PW, Lesser ER, et al. Safety Update: COVID-19 Convalescent Plasma in 20,000 Hospitalized Patients. Mayo Clin Proc. 2020 Sep;95(9) 1888–1897. doi:10.1016/j.mayocp.2020.06.028. PMID: 32861333; PMCID: PMC7368917.

24. Liu STH, Lin HM, Baine I, Wajnberg A, Gumprecht JP, Rahman F, Rodriguez D, Tandon P, Bassily-Marcus A, Bander J, Sanky C, Dupper A, Zheng A, Nguyen FT, Amanat F, Stadlbauer D, Altman DR, Chen BK, Krammer F, Mendu DR, Firpo-Betancourt A, Levin MA, Bagiella E, Casadevall A, Cordon-Cardo C, Jhang JS, Arinsburg SA, Reich DL, Aberg JA, Bouvier NM. Convalescent plasma treatment of severe COVID-19: a propensity score-matched control study. Nat Med. 2020. 10.1038/s41591-020-1088-9 Epub ahead of print.

25. Rogers R, Shehadeh F, Mylona EK, Rich J, Neill M, Touzard-Romo F, et al. Convalescent plasma for patients with severe COVID-19: a matched cohort study. Clinical Infectious Diseases [Internet]. 2020; Disponible en: https://doi.org/10.1093/cid/ciaa1548

26. Plasma trials [Internet]. COVID-19 research and trials - NHS Blood and Transplant. [citado 8 de diciembre de 2020].Disponible en: /covid-19-research/research-andtrials/plasmatrials/

